# Antibody responses to SARS-CoV-2 vaccination in patients with acute leukaemia and high risk MDS on active anti-cancer therapies

**DOI:** 10.1101/2022.04.05.22273371

**Authors:** W.Y. Chan, C. Zhu, E. Sanchez, R. Gupta, A.K Fielding, A. Khwaja, E.M. Payne, J. O’Nions

## Abstract

Patients with haematological malignancies, such as acute leukaemia and high-risk MDS (HR-MDS), have significantly increased mortality and morbidity from COVID-19. However vaccine efficacy in these patients and the impact of systemic anti-cancer therapy (SACT) on vaccine response remains to be fully established. SARS-CoV-2 antibody responses in 53 patients with ALL, AML or HR-MDS receiving SACT were characterised following two doses of either BNT162b2 or ChAdOx1nCoV-19. All patients were tested for anti-S antibodies after 2 doses, 60% after the first dose and anti-N antibody testing was performed on 46 patients (87%). Seropositivity rates after 2 vaccine doses were 95% in AML/HR-MDS patients and 79% in ALL. After stratification by prior SARS-CoV-2 infection, naïve patients with AML/HR-MDS had higher seroconversion rates and median anti-S antibody titres compared to ALL (median 291U/mL versus 5.06U/mL), and significant increases in anti-S titres with consecutive vaccine doses, not seen in ALL. No difference was seen in serological response between patients receiving intensive chemotherapy or non-intensive therapies (HMA) but significantly reduced titres were present in AML/HR-MDS patients who received venetoclax-based regimens compared to other therapies. All ALL patients received intensive chemotherapy, with no further impact of anti-CD20 immunotherapy on serological response. Understanding the impact of disease subtypes and therapy on vaccine response is essential to enable decisions on modifying or delaying treatment in the context of either SARS-CoV-2 infection or vaccination.

## Introduction

SARS-CoV-2 vaccination represents an important measure to protect the population from COVID-19, showing excellent rates of seroconversion and efficacy in preventing severe disease^1,2^. However, vaccine efficacy remains to be fully-established in patients with haematological malignancy who have significantly increased mortality and morbidity from COVID-19^3,4,5^. Variable humoral responses to SARS-CoV-2 vaccination have been reported, with the lowest seroconversion rates in patients with B-cell malignancies and those receiving systemic anti-cancer therapy (SACT)^6,7,8,9^. Encouraging seroconversion rates in patients with acute lymphoblastic leukaemia (ALL), acute myeloid leukaemia (AML) and HR-MDS have been reported within larger cohort studies of patients with a variety of haematological cancers^7,10,11,12^. However, the impact of SACT on vaccine responses in patients with ALL, AML and high-risk myelodysplastic syndrome (HR-MDS) remains unclear, but has important implications for clinical management. Here we have addressed this and report SARS-CoV-2 antibody responses following vaccination in patients with ALL, AML or HR-MDS receiving SACT.

## Methods

Demographics, SACT history and laboratory parameters were collected from electronic health records for patients with acute leukaemias and HR-MDS receiving or recently completed SACT, who had two doses of SARS-CoV-2 vaccine (BNT162b2 or ChAdOx1nCoV-19) between December 2020 and July 2021 (with 8 to 12 weeks between doses as per UK vaccination programme). Serological testing was performed using the Roche Elecsys anti-SARS-CoV-2 enzyme immunoassays. All patients were consented for excess serum to be stored and used as part of the “UCL Biobank for Studying Health and Disease, Haematology Project” (ref.NC10.13).

## Results

Fifty three patients (62% AML, 11% HR-MDS, 26% ALL), underwent serological testing after receiving 2 vaccine doses (72% BNT162b2, 15% ChAdOx1nCoV-19, 13% unknown), Table 1. Median age was 53 (range 18-76) and all received or had recently completed SACT at time of vaccination; 21 patients (39.6%) received intensive chemotherapy, 37.7% venetoclax-combination therapy (with azacitidine, low-dose cytarabine or gilteritinib), 16.9% non-intensive (azacitidine) and 5.6% B-cell directed immunotherapy (blinatumomab, CD19-CART). Fourty-six (87%) underwent testing for anti-N antibodies, identifying 11 (24%) showing previous SARS-CoV-2 infection. All patients were tested for anti-S antibodies after 2 doses (median 43 days post-dose) and 60% after first dose (median 37.5 days). A trend towards higher rates of seropositivity was seen in AML and HR-MDS compared to ALL after 2 doses (95% compared to 79%, p=0.06, Figure 1A). The seropositivity rate after dose one in all patients was 75% (median anti-S titre 7.275U/ml [IQR 0.64-184]), rising to 91% following dose two (median 249U/ml [IQR 24.95-1721]), Figure 1B. There was no significant difference in anti-S seropositivity rate or median antibody titre after dose one between patients with AML/HR-MDS and ALL (74% vs 78% seropositive, median titres 5.90U/ml [IQR 0.58-56.70] versus 11.4U/ml [IQR 0.61-1380]). However, patients with AML/HR-MDS showed significantly increased titres following dose two (median 333U/mL [IQR 86.60-1971], p=0.0005, 95% seropositive), compared to ALL (median titre 8.40 [IQR 2.34-669], p=0.38, 79% seropositive).

**Table 1:** Patient demographics, disease and treatment characteristics in the overall cohort and by disease subgroup.

| Characteristics | Total<br>*n*=53 | Negative<br>baseline<br>*n*=35 | Positive<br>baseline<br>*n*=11 | Myeloid (AML<br>+ HR-MDS)<br>*n*=39 | Lymphoid<br>*n*=14 |
| --- | --- | --- | --- | --- | --- |
| Gender (% male) | 30 (57) | 18 (51) | 6 (55) | 21 (54) | 9 (64) |
| Median age [range] | 53 [18-76] | 52 [18-76] | 43 [22-73] | 63 [21-76] | 37 [18-57] |
| Diagnosis (%): |  |  |  |  |  |
| AML/Myeloid Sarcoma | 28 (53) | 19 (54) | 4 (36) | 28 (72) | - |
| APML | 5 (9) | 3 (9) | 2 (18) | 5 (13) | - |
| HR MDS | 6 (11) | 4 (11) | 1 (9) | 6 (15) | - |
| B-ALL | 10 (19) | 7 (20) | 3 (27) | - | 10 (71) |
| T-ALL | 4 (8) | 2 (6) | 1 (9) | - | 4 (29) |
| SARS-CoV-2 infection*(%) | 11/46 (24) | - | - | 7/33 (21) | 4/13 (31) |
| Treatment (% total patients): |  |  |  |  |  |
| Intensive AML chemotherapy | 10 (19) | 7 (20) | 3 (27) | 10 (26) | - |
| Venetoclax based regimens | 20 (38) | 12 (34) | 3 (27) | 20 (51) | - |
| - Ven & Aza | 16 | 10 | 2 | 16 | - |
| - Ven & LDAC | 2 | 1 | 0 | 2 | - |
| - Ven & Gilt | 1 | 1 | 0 | 1 | - |
| - Ven, Gilt & Aza | 1 | 0 | 1 | 1 | - |
| Non intensive AML therapy | 9 (17) | 7 (20) | 1 (10) | 9 (23) | - |
| UKALL chemotherapy | 11 (21) | 9 (26) | 2 (20) | - | 11 (79) |
| - Plus Rituximab | 5 | 3 | 2 | - | 5 |
| Other B cell directed therapy | 3 (6) | 1 (3) | 2 (20) | - | 3 (21) |
| - Blinatumumab | 2 | 1 | 1 | - | 2 |
| - CAR-T | 1 | 0 | 1 | - | 1 |
| Seropositive**, 1 dose (%) | 24/32 (75) | 13/18 (72) | 10/11 (91) | 17/23 (74) | 7/9 (78) |
| Seropositive**, 2 doses (%) | 48/53 (91) | 32/35 (91) | 10/11 (91) | 37/39 (95) | 11/14 (79) |
| Seroconversion*** post 2 doses (%) | 32/35 (91) | 32/35 (91) | - | 25/26 (96) | 7/9 (78) |
| Vaccine type (%): |  |  |  |  |  |
| BNT162b2 | 38 (72) | 23 (66) | 11 (100) | 26 (67) | 12 (86) |
| ChAdOx1 nCoV-19 | 8 (15) | 6 (17) | 0 (0) | 8 (21) | 0 (0) |
| unknown | 7 (13) | 6 (17) | 0 (0) | 5 (13) | 2 (14) |
| Median time (days) from first dose to serology [range] | 37.5 [24-86] | 31 [24-79] | 44 [24-79] | 39 [24-79] | 36 [26-86] |
| Time (days) from second dose to serology [range] | 43 [13-133] | 41 [13-133] | 45 [15-72] | 40 [13-133] | 48 [15-84] |
| Median titres post first dose in all patients U/mL (IQR) | 7.275 U/mL (0.64-184) | 4.42 U/mL (0.4-17.10) | 2500 U/mL (259-2500) | 5.90 U/mL (0.58-56.70) | 11.40 U/mL (0.61-1380) |
| Median titres post first dose in patients with negative baseline U/mL (IQR) | N/A | 4.42 U/mL (0.4-17.10) | N/A | 5.90 U/mL (0.62-38.35) | 0.82 U/mL (0.4-9.64) |
| Median titres post second dose in all patients U/mL (IQR) | 249 U/mL (24.95-1721) | 132 U/mL (7.5-931) | 2500 U/mL (417-2500) | 333 U/mL (86.80-1971) | 8.40 U/mL (2.34-669) |
| Median titres post second dose in patients with negative baseline U/mL (IQR) | N/A | 132 U/mL (7.5-931) | N/A | 235U/mL (82.15-1670) | 5.06U/mL (1.69-175.6) |

\*SARS-CoV-2 infection defined by presence of anti-N (nucleocapsid) antibodies.
\*\*SARS-CoV-2 seropositive defined by presence of anti-S (Spike) antibodies.
\*\*\*Seroconversion defined by the detection of anti-S antibodies in patients who had previously undetectable anti-S antibodies.
Definitions: AML, acute myeloid leukaemia; HR-MDS, high risk MDS; B-ALL, B-lymphoblastic leukaemia; T-ALL, T lymphoblastic leukaemia; Ven, venetoclax; Aza, azacitidine; LDAC, low dose cytarabine; Gilt, gilteritinib; UKALL, (UK ALL chemotherapy 2011, 2014 protocols [anthracycline, vincristine, cytarabine, cyclophosphamide, peg-asparaginase, dexamethasone])

**Figure 1:**
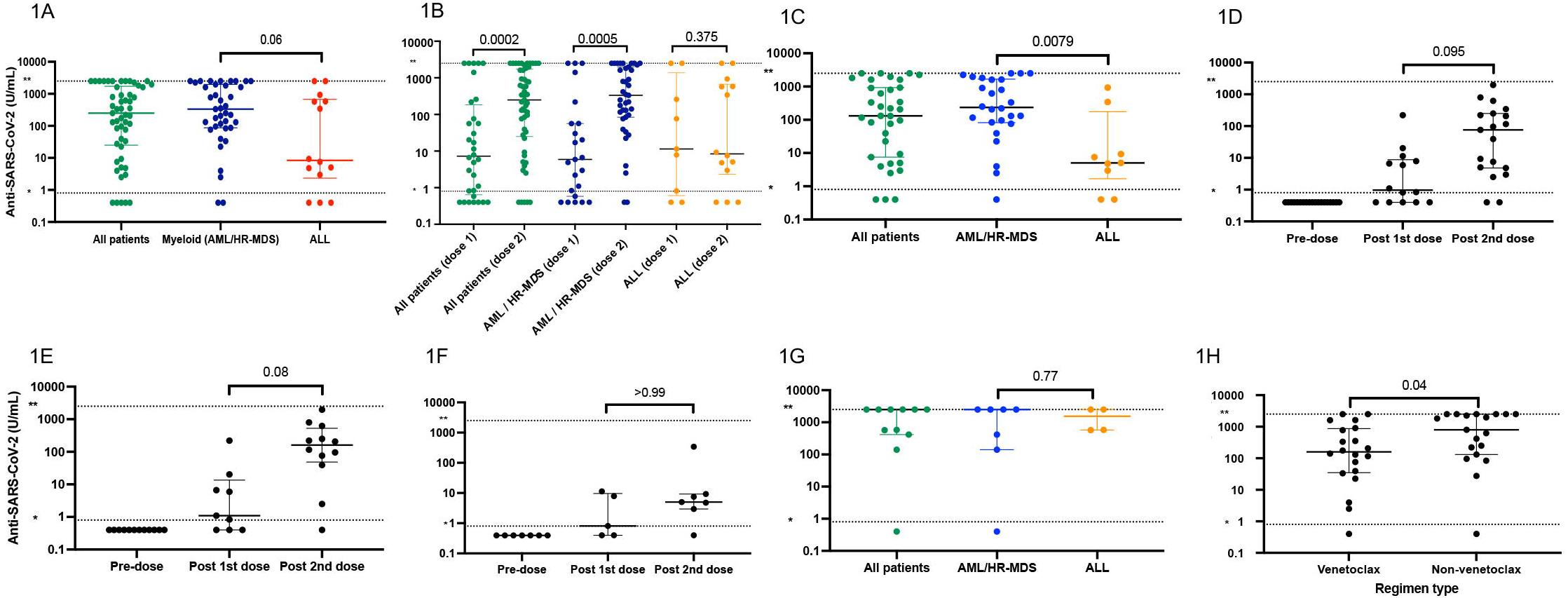
Serological responses in patients with acute leukaemia and HR-MDS after SARS-CoV-2 vaccination. All figures are presented with a Log10 scale on the y-axis. A seropositive result is defined as ≥0.8U/mL, upper limit of the assay >2500U/ml *\*lower limit of assay, **upper limit of assay*. **1A**: Seropostivity for anti-S antibodies in all patients following two doses of SARS-CoV-2 vaccine, categorized by disease subtype. **1B:** Anti-S antibody titres following the first and second vaccine doses by disease category. **1C:** Serological response to two vaccination doses in patients with no previous SARS-CoV-2 infection. **1D:** Seroconversion rates in patients with no previous SARS-CoV-2 infection, after one and two doses of vaccine (Paired pre dose, post first dose and second dose); in patients with AML or HR-MDS **(1E)** and in patients with ALL **(1F). 1G:** Serological response in patients with previous SARS-CoV-2 infection categorized by disease subtypes. **1H:**Serological response following two doses of vaccine in AML/HR-MDS treated with venetoclax-based regimens.

To define vaccine-induced seroconversion, we analysed responses in patients with no prior SARS-CoV-2 infection and negative anti-N serology (Figure 1C, Figure S1A). Anti-S seroconversion was 91% with median titre of 132U/mL (IQR 7.5-931). Seroconversion rates and median titres in SARS-CoV-2 naïve patients remained higher in the myeloid compared to lymphoid cohort (median 235U/mL [IQR 82.15-1670] versus 5.06U/mL [1.69-175.6], p=0.0079). A marked increase in titres was seen following consecutive vaccinations in the AML/HR-MDS cohort (medians post dose one and two of 1.1U/mL and 95.8U/mL, p=0.08), but only minimal increase in ALL (0.817U/ml to 4.80U/mL, p>0.99) (Figures 1D, E, F, Figures S1B, C). Previous SARS-CoV-2 infection was associated with higher antibody titres after two vaccinations in patients with AML/HR-MDS (median 2500U/mL [IQR 141-2500]) compared to ALL (median 1541U/mL [IQR 574.5-2500], although this did not reach significance (p=0.77) (Figure 1G, Figures S1D,E). This is consistent with reports of higher overall post-vaccination antibody titres in individuals with prior natural infection^13,14^.

Next we investigated the impact of SACT type on serological responses. No significant difference in seropositivity or antibody titres was seen in AML/HR-MDS patients receiving intensive (30% patients) compared to non-intensive chemotherapy (azacitidine, 23%); however, anti-S titres were significantly reduced in patients on venetoclax-based regimens (51% of myeloid patients, median 158.5U/mL [IQR 34.85-873], p=0.04). This was independent of previous SARS-CoV-2 infection (Figure 1H, Figure S2A). The majority of ALL patients (79%) received intensive treatment (as per current UKALL protocols) and had low anti-S titres post vaccination (Figure 1A,B,C,F). Anti-B cell directed therapy is associated with poor serological response to SARS-CoV-2 vaccination. Five ALL patients received Rituximab but no effect was seen on anti-S response, albeit in small numbers (Figure S2B,C).

Here we report the majority of patients with acute leukaemias and HR-MDS develop detectable anti-S antibodies following two doses of SARS-CoV-2 vaccine, with seropositivity rates of 91%. SARS-CoV-2 naïve patients with AML/HR-MDS showed significantly increased antibody responses following consecutive vaccine doses, whereas ALL patients tended towards very low titres with a minimal increase, and it remains to be established whether this is sufficient to confer protective immunity. While patients with acute leukaemias can generate robust serological responses to natural infection, this was not the case for all patients following vaccination, highlighting the importance of measuring antibody titres, not just seropositivity, and to consider prior SARS-CoV-2 infection influencing vaccine responses in this high-risk group. Understanding the impact of SACT on SARS-CoV-2 vaccine responses is essential to guide decisions on treatment choice and timing. Reduced serological responses in patients receiving B-cell directed therapy and small molecule inhibitors (particularly BTKi, venetoclax, anti-CD20 and anti-CD19 immunotherapies) have been reported in mature B-cell neoplasms and myeloma, but not acute leukaemias^6,9,11,15^. While we found no significant difference in antibody titres or seroconversion in AML/HR-MDS patients receiving intensive or non-intensive therapy, patients receiving venetoclax-based regimens showed reduced antibody levels. Notably, ALL with intensive chemotherapy displayed almost uniformly low antibody titres (<10U/ml) after two vaccine doses, regardless of additional B-cell directed therapy. We propose that further work to define correlates of vaccine protection (humoral and T cell responses), the impact of SACT regimens on the magnitude/duration of responses, and the value of additional booster doses will have clear implications for this vulnerable group and should be priority questions for larger prospective studies.

## Data Availability

All data produced in the present work are contained in the manuscript

## Contributions

JO conceived of the study, performed data collection, data analysis, literature search, and manuscript writing and revision; WC data collection, data analysis, manuscript writing, and revision; CZ performed data collection, manuscript review and revision; EP conceived of study, manuscript review and revision; ES data collection, manuscript review and revision; AF, AK, RG, manuscript review and revision.

## Declaration of interest

All authors declare no conflicts of interest.

## Acknowledgements

We are extremely grateful to all the patients who participated in this study and to the NHS staff that provided their clinical care. EMP is supported by a CRUK Advanced Clinician Scientist Fellowship (Grant No. A24873). RG acknowledges funding from Cure Cancer@ UCL.

## Notes

### Competing Interest Statement

The authors have declared no competing interest.

### Author Declarations

All clinical information was recorded and blood samples taken as routine standard of care. Patients were consented to allow any excess serum to be stored and used as part of the UCL Biobank for Studying Health and Disease, Haematology Project, reference no NC10.13, approved by the Leeds (East) Research Ethics Committee, UK.

