## supplemental figures 1 and 2 for "Antibody responses to SARS-CoV-2 vaccination in patients with acute leukaemia and high risk MDS on active anti-cancer therapies"

**Supplementary Figure 1: Trajectories of anti-SARS-CoV-2 Spike protein (anti-S) IgG titres in individual patients following first and second doses of vaccine, stratified by disease subtype.**

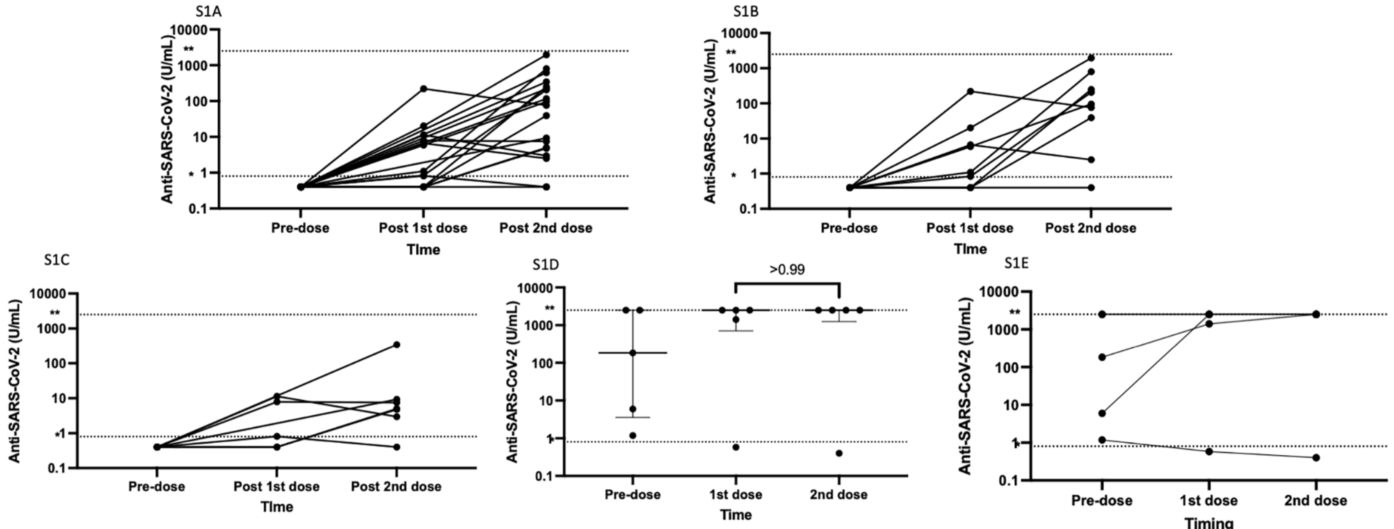


**Figure legend: All figures are presented with a Log10 scale on the y-axis.**
**lower limit of assay, **upper limit of assay, HR-MDS High Risk MDS*
**S1A:** Line graph seroconversion rates in patients with no previous SARS-CoV-2 infection, after one and two doses of vaccine (Paired pre dose, post first dose and second dose) in all patients; in patients with AML or HR-MDS **(S1B)** and in patients with ALL **(S1C)**
**S1D:** Seroconversion rates in all patients with previous SARS-CoV-2 infection, after one and two doses of vaccine (Paired pre dose, post first dose and second dose); displayed as a line graph **(S1E)**

### Supplementary Figure 2: Serological responses in patients with Acute leukaemia stratified by therapy

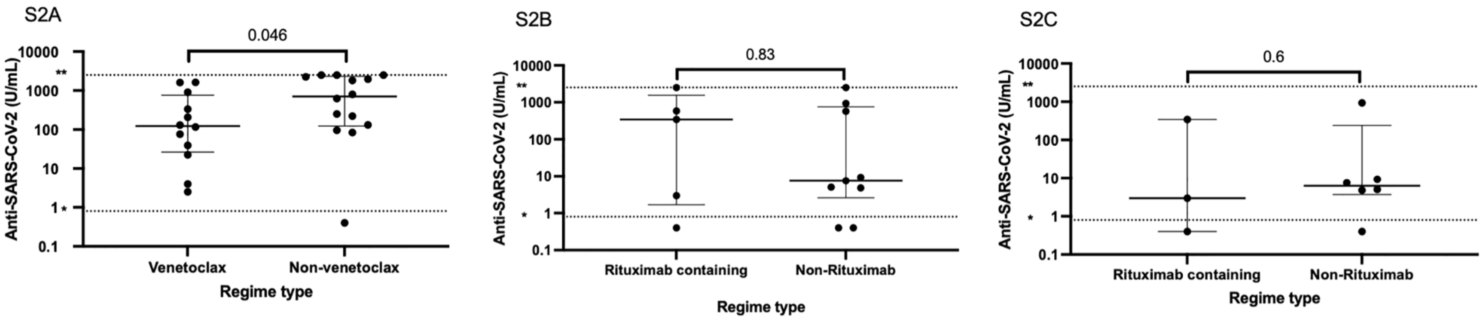


**Figure legend: All figures are presented with a Log10 scale on the y-axis.**
**lower limit of assay, **upper limit of assay, HR-MDS High Risk MDS*
**S2A:** Serological response following two doses of vaccine in AML/HR-MDS treated with venetoclax-based regimens and no evidence of previous SARS-CoV-2 infection
**S2B:** Serological response following two doses of vaccine in patients treated with Rituximab and non-Rituximab containing regimes: Median 342 U/mL (IQR 1.69-1541) vs Median 7.5U/mL (IQR 2.6-751.5) p=0.83
**S2C:** Serological response following two doses of vaccine in patients treated with Rituximab and non-rituximab containing regimes (Baseline COVID –ve only): Median 2.980 (IQR 0.4-342) vs Median 6.280 U/mL (IQR 3.7-239.7) p=0.6
